# Child poverty and declining measles, mumps and rubella (MMR) vaccination in England, 2015 to 2024. A longitudinal ecological study at local area level

**DOI:** 10.64898/2026.03.10.26348016

**Authors:** Yu Wei Chua, Luke Munford, Olivia Pearce, Helen Skirrow, Miriam Taegtmeyer, Neil French, Matthew Ashton, Daniel Hungerford, David Taylor-Robinson

## Abstract

**Objective:** To assess the contribution of changing child poverty rates to trends in measles, mumps and rubella (MMR) vaccination.

**Design:** Longitudinal area-level analysis using within-between models to assess the association of increases in child poverty within-areas on MMR vaccination

**Setting:** 148 upper-tier local authorities in England from 2015 and 2024.

**Participants:** Children aged 5 years or younger eligible for MMR vaccination in England between 2015 and 2024. 6,468,620 children aged 24 months were included in the study and 6,907,640 aged 5 years.

**Exposures:** Child poverty rates for each upper-tier local authority, measured as the percentage of children aged 0 to 15 living in households below 60% of Organisation for Economic Co-operation and Development (OECD) median, before housing costs.

**Outcome Measures:** MMR 1^st^ and 2^nd^ dose uptake rates by 24 months 5 years of age respectively, at upper-tier local authority.

**Results:** Over the study period, MMR 1^st^ dose fell by 4.0 percentage-points (%) (range: −20.8 to 7.7) and MMR 2^nd^ dose by 4.9% (range: −23.4 to 10.1) while child poverty rose by 5.6% (range: 0.2 to 13.9) on average. A 1 percentage-point (%) increase in child poverty was associated with a 0.17% [95%CI: −0.29; −0.06] fall in MMR 1^st^ dose rates and a 0.26% [95%CI: −0.42; −0.10] fall in MMR 2^nd^ dose rates.

**Conclusion:** Rising child poverty rates have contributed to a decrease in MMR vaccination in children in England. Action to reduce child poverty is needed to improve childhood vaccination uptake alongside policies and interventions specific to vaccination and infectious disease prevention.

**Summary box:** *What is already known on this topic:* Inequalities in childhood vaccination uptake in England are stark and have widened, especially for MMR vaccination. Child poverty in England has increased and is associated with rising inequalities in multiple domains of children’s health but impacts on inequalities in vaccination uptake are unclear.

*What this study adds:* A 1 percentage-point increase in child poverty was associated with a 0.17 percentage point fall in uptake of MMR1 and a 0.26 percentage point fall in MMR2 between 2015 and 2024. Reducing child poverty is likely to increase vaccine uptake and reduce the burden of vaccine preventable diseases in England.

---

Childhood vaccine uptake in England has been declining since the late 2010s, with the COVID-19 pandemic exacerbating this decline, and widening inequalities between advantaged and disadvantaged socioeconomic groups (1). This is particularly evident for measles, mumps, and rubella vaccination (MMR), with the largest measles outbreaks since 2012, including 2,911 confirmed cases in 2024 (2). England lost its measles elimination status in 2019, regaining it temporarily during the COVID-19 pandemic due to reduced transmission of all other infectious diseases (3). Vaccine preventable diseases cost the National Health Service (NHS) £6 billion annually for secondary care services alone (4). Large outbreaks of measles, which disproportionately affect those living in poverty, can cost society more than 20 times the amount needed for preventative vaccination (5). Despite a comprehensive immunisation schedule, those most at risk of severe disease and long-term health complications can miss out on vaccination in the United Kingdom (UK), and uptake is below levels needed for herd immunity (3). Geographically, the North of England, the Midlands, and London are most affected by these inequalities in vaccine uptake.

These widening inequalities in vaccine uptake occur against a backdrop of government austerity policies since 2010, which have led to cuts in the NHS, public services, local government, and welfare support (6,7). Since 2015, funding for health visitors has declined by 27%; early years services such as Sure Start saw a two-thirds reduction in investment between 2010 and 2022, leading to the closure of 1,340 centres (8). These policies, including removal of child benefits for the third child or additional children, changes to housing benefit, and the introduction of Universal Credit have disproportionately affected low-income and vulnerable populations. The covid-19 pandemic, the global energy crisis, and Brexit have worsened the ongoing cost-of-living-crisis in the UK and increased child poverty(9), leading to poor health outcomes, including higher rates of infant mortality and children entering care (10,11). In 2024, an estimated 31% of children (4.5 million) in the UK live in poverty, despite most of these children residing in working households (12). Child poverty is most prevalent in the Northwest of England, West Midlands, and inner London, and is disproportionally higher in ethnic minority households.

Child poverty and low vaccine uptake independently contribute to poor child health outcomes such as infant mortality, so understanding the relationship between child poverty and childhood vaccination uptake is crucial for informing policy changes that address preventable health inequalities. The Government’s Child Poverty Strategy ends policies such as the two-child cap to child benefit and is estimated to take half a million children out of poverty and remove a further 700,000 from deep poverty (13). Such changes may increase vaccination uptake and improve health outcomes, especially in deprived areas. In this study, we use area-level data from English local authorities (LA) to examine the association between child poverty rates on time trends and area-level variation in Measles, Mumps, and Rubella vaccination (MMR) 1^st^ dose by 24 months and MMR 2^nd^ dose by 5 years. Our primary aim was to assess how **changes in child poverty rates over time** have impacted MMR rates, between 2015 and 2024. As a secondary aim, we describe how **area-level differences in child poverty rates** explain variation in MMR vaccination rates between local authorities over the study period.

## Methods

### Study design and setting

This was a longitudinal, ecological study of upper-tier local authorities in England, UK. Upper-tier local authorities are administrative regions responsible for delivering a range of local services such as education and social care (14).

### Eligibility criteria

Local authorities with reconcilable boundaries which had data available on MMR immunisation and child poverty were eligible. Due to small local authority populations, data from Rutland, Isles of Scilly, and City of London were combined respectively with Leicestershire, Cornwall, and Hackney. To deal with boundary changes, we combined West and North Northamptonshire and excluded data from Bournemouth and data from Poole from 2014 to 2019 (as Bournemouth, Christchurch and Poole was formed in 2020). Westmorland and Furness, formed in 2023, had no vaccination data. 148 local authorities were eligible for inclusion with data on both vaccination and child poverty data available from 2014/15 to 2023/24 (hereafter 2015 to 2024).

### Data sources and measurement

Outcome data on MMR immunisation was obtained from the Childhood Vaccination Coverage Statistics published by NHS England/UK Health Security Agency (UKHSA) (15), available annually from 2014 to 2024. We calculated the percentage coverage of MMR 1^st^ dose at 24 months (MMR1 24M) and MMR 2^nd^ dose at 5 years (MMR2 5Y), based on data on the number of number of children eligible and children receiving the vaccine.

Exposure data on child poverty was obtained from the Children in Low Income Families dataset, available annually from 2015 to 2024 (16). Child poverty was defined as the percentage of children aged 0 to 15 living in households below 60% of Organisation for Economic Co-operation and Development (OECD) median, before housing costs. Since data was available at the Lower Tier Local Authority level, we calculate child poverty rates for each upper-tier local authority by summing the numbers for each lower-tier authority within each upper-tier local authority, and dividing by Office for National Statistics population estimates of children 0 to 15 years for each LA and year (17).

Covariates were considered based on a logic model on factors that could influence child poverty and vaccination rates (Figure S1). Time period was defined as a categorical variable to control for annual trends and national time shocks (18). Controlling for time period as dummy variables controls for common time shocks experienced by all local authorities. We controlled for pressure on health services resulting from covid-19 as a confounder, proxied using covid-19 excess mortality published by the Office for Health Improvement and Disparities. Local authorities experienced varying levels of COVID-19–related strain on health and public health services (19), which may have affected both the delivery of vaccination services and vaccine uptake (20). We included local authority minority ethnic composition and educational qualifications based on the 2021 UK census, defined respectively as the percentage of children aged 0 to 15 of White British/Irish ethnicity (21), and rank of the highest level of educational qualification index score (as defined by the Office for National Statistics). We coded the education rank such that a higher rank represented lower educational qualifications relative to other local authorities. We also included annual standardised mean maternal age, available from 2013 to 2024, from the Office for National Statistics Live Births in England and Wales: birth rates down to local authority dataset (22).

### Statistical methods

#### Statistical analyses were carried out in R (V4.3.2)

We visualised annual time trends using mean and 95% confidence intervals. We obtained the overall difference between 2015 and 2024 and visualised geographical differences using (*sf* package), mapping values onto Office for National Statistics Counties and Unitary Authorities 2022 vector boundaries. We obtained the annual lagged differences and averaged this over the period 2015 to 2024. We visualised the time-averaged annual lagged difference of child poverty and MMR1 24M and MMR2 5Y using scatter plots.

We fitted within-between models of MMR1 24M and MMR2 5Y against child poverty (23), controlling for Year fixed effects (*lmer, lme4* package). The “Within-area” estimates address our primary research question, capturing the average association of annual changes in child poverty and annual changes in MMR rates. We focus on the within-area estimates as they are more robust to confounding by a range of local authority characteristics that remain relatively constant over time. The “between-area’ estimates capture the observed association of differences in child poverty and vaccination rates between local authorities and address our secondary research question. Between-area estimates can still be influenced by a range of characteristics correlated with poverty, that differ between local authorities. To allow the associations of child poverty to vary before versus after covid-19, we considered interactions within-area and between-area child poverty terms with a covid-19 indicator variable (0: 2015-2019, 1: 2020-2024), assessed using comparison of model fit (Akaike and Bayesian Information Criterion and likelihood ratio tests).

We fitted unadjusted models, and models adjusted for confounders (time-invariant minority ethnic composition, education rank, and covid-19 excess mortality, and time-varying maternal age). Where interaction with covid-19 was present, we obtained simple slopes for the association of child poverty on MMR rates (i.e., representing pre- and post-covid-19 associations) (*emmeans* and *ggeffects* package). We centred year so that between-area effects are estimated in 2019, the median year. We scaled child poverty so that main effects capture a 1 percentage-point increase or difference in child poverty rates. We scaled and centred covariates (see Appendix), so parameter estimates capture associations for the “average” local authority (the mean over area and time) of child poverty and maternal age, the mean (over area) of ethnicity and excess mortality, and 50^th^ percentile of education rank).

We calculate the observed total number of children who did not receive the MMR 2^nd^ dose by 5 years, and the numbers expected had MMR 2^nd^ dose coverage remained at 2015 levels. We calculated the change in number of 5-year-old children not fully protected by the MMR vaccination course attributable to the cumulative changes in child poverty: first, within each area, we calculated the change in child poverty in subsequent years relative to 2015; second we calculated the number of unvaccinated children by applying the within-area estimates of child poverty on MMR 2^nd^ dose rates to the change in child poverty and the annual population eligible for the 2^nd^ dose vaccination, and summed across local authorities.

In sensitivity analyses, first, we repeated the analysis to assess the lagged association of child poverty on MMR1 24M and MMR2 5Y rates 1 year later, for MMR rates measured between 2016 to 2024.

Second, we re-ran the models using fixed effect models, where between-area variability is subsumed in the area-level intercept, to test the robustness to of the estimates to different model specification. Since the effect of between-area child poverty differs by period in the main analysis, we stratified the fixed effect models by pre- and post-covid-19 periods, as the area-level intercept (used for de-meaning variables in the fixed effects specification) would be mis-specified if assumed to be constant over the whole period. Finally, we considered first-order interactions of child poverty and ethnicity, education, and maternal age - to fully describe how “between-area” characteristics influenced geographic variations in MMR trends (see Appendix, Table S1 and S2 for further methodological details).

### Patient public involvement

No patients or members of the public were directly involved in this research. But the research questions were informed by prior patient public involvement and engagement activities. As part of the IMPRINT (Immunising PRegnant women and INfanTs) network: https://www.imprint-network.co.uk/news/public-engagement-project-completed-2), multi-national consultation groups and engagement activities were held with parents and carers on immunisation topics including benefits, concerns and barriers, and research priorities. Locally, community innovation teams provided insights from community members for shaping research priorities for work on reducing inequalities in MMR vaccine uptake, through the UKRI funded ReCITE (https://www.lstmed.ac.uk/recite) project. The research questions were also informed by our engagement with children and young people through the Health Equity North/Child of the North initiative (https://www.healthequitynorth.co.uk/child-of-the-north/), which highlighted a key focus on tackling child poverty. The findings from this study are being shared with patients and communities through the above programmes, but also with public health organisations and presented at regional (e.g. Health Equity Liverpool Community Engagement Event) and national events, with health, lay, and government representation.

## Results

### Sample characteristics

148 (100%) local authorities with data between 2015 to 2024 on child poverty and MMR 1^st^ dose by 24 months and MMR 2^nd^ dose by 5 years. Data from Bournemouth, Christchurch and Poole was available from 2020 to 2024. Data on all covariates were complete. Across 2015 to 2024, a total of 6,468,620 children were eligible for MMR 1^st^ dose and 6,907,640 for MMR 2^nd^ dose.

Average child poverty rates increased from 14.7 percentage-points (%) [95%CI: 13.4; 15.9] in 2015 to 20.3% [18.5; 22.0] in 2024. Average MMR 1^st^ dose and MMR 2^nd^ dose rates fell steadily from 2015 (MMR1 24M: 92.4% [91.8; 93.1]; MMR2 5Y: 88.8% [87.9; 89.8]) to 2024 (MMR1 24M: 88.8% [88.0; 89.7]; MMR2 5Y: 83.6% [82.4; 84.8]) (see Figure 1 and Table S3). Over the 10-year-period, a total of 608030 children did not receive MMR 1^st^ dose by 2 years, and 928066 children did not receive MMR 2^nd^ dose by 5 years.

From 2015 to 2024, child poverty within a local authority increased on average by 5.6 percentage-points (%) (range: 0.2 to 13.9); MMR1 24M fell by 4.0% (range: −20.8 to 7.7); and MMR2 5Y fell by 4.9% (range: −23.4 to 10.1). Of 29 local authorities with the largest increase in child poverty rates (Top quintile: 9 to 13.9%), 12 (41.4%) had the largest fall in MMR1 24M rates (Bottom quintile: −20.6 to −6.9%) and 16 (55.2%) had the largest fall in MMR2 5Y rates (Bottom quintile: −28.6 to - 8.5%). The areas with the largest rise in child poverty, alongside the largest fall in both MMR1 24M and MMR2 5Y were Birmingham, Bradford, Hackney and City of London, Knowsley, Leeds, Leicester, Liverpool, Nottingham, Oldham, Stoke-on-Trent, and Walsall (Figure 4).

**Figure 2.**
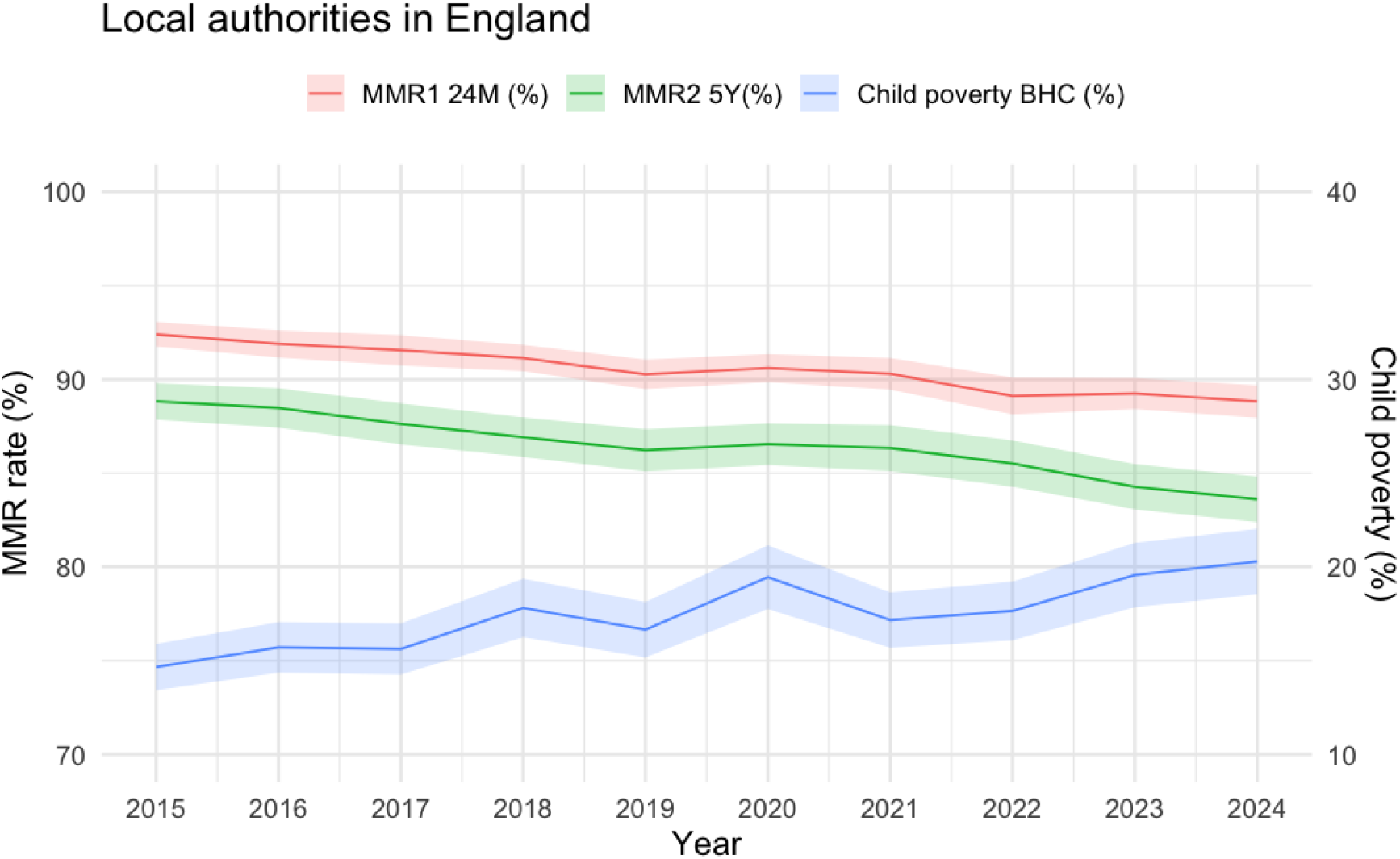
Average rates across local authorities in England in child poverty before housing costs (BHC), MMR 1^st^ dose at 24 months (MMR1 24M) and MMR 2^nd^ dose at 5 years (MMR2 5Y) in the years 2015 to 2024

**Figure 3.**
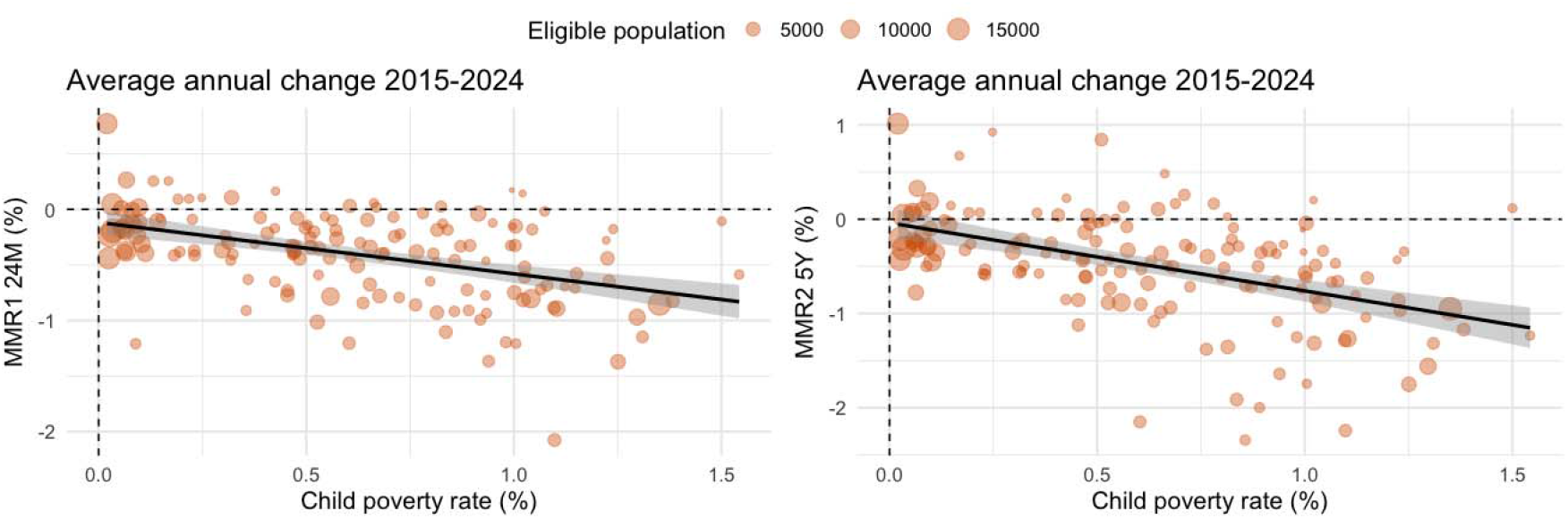
Descriptive scatterplots of local authority average annual change (from 2015 to 2024) in child poverty and MMR rates (Left: MMR1 24M; Right: MMR2 5Y)

**Figure 4.**
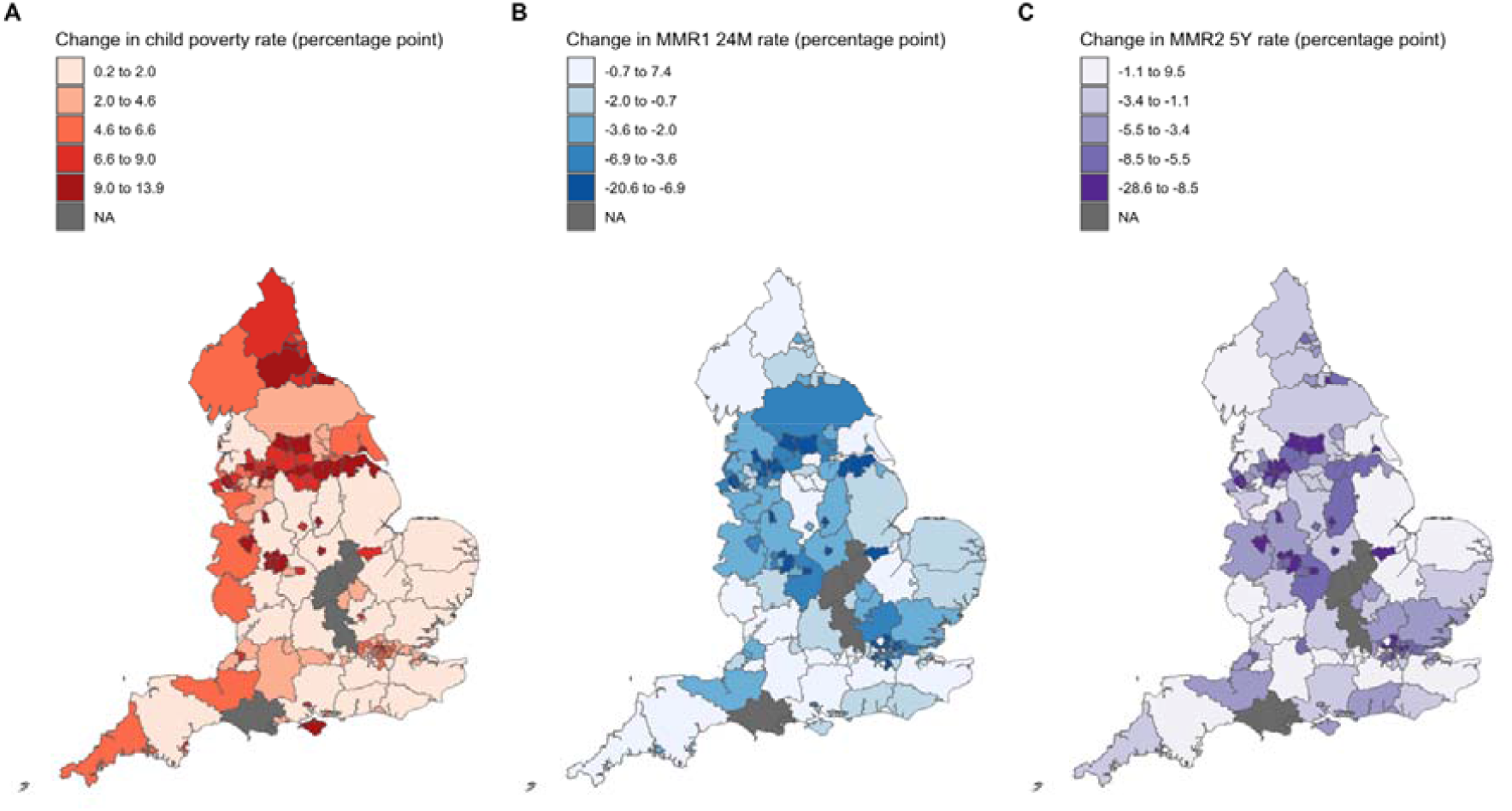
Map of England showing the overall change between 2015 to 2024 in rates of child poverty (panel A), MMR1 24M (panel B) and MMR2 5Y (panel C), coloured by quintiles. Greyed areas show local authorities with small populations combined with another local authority or areas with missing data in 2015 or 2024 due to boundary changes (i.e., West and North Northamptonshire; Bournemouth, Christchurch and Poole; Westmorland and Furness).

### Within-area association of change in child poverty on MMR vaccination rates

In confounder adjusted models, a 1 percentage-point increase in child poverty rate within a local authority was associated, on average, with a −0.17% [-0.29; −0.06] (p=0.003) fall in MMR1 24M rates and a larger fall, −0.26% [-0.42; −0.10] (p<0.001), in MMR2 5Y rates (Table 1). Likelihood ratio tests did not find evidence of an interaction between the covid-19 dummy variable and the within-area child poverty term ( χ^2^(1) MMR1 24M = 0.034, p=0.855; MMR2 5Y=0.162; p=0.682), so this term was not included in either model. Adjusted and unadjusted within-area child poverty were essentially same (see Table S4 and S5).

**Table 1.**
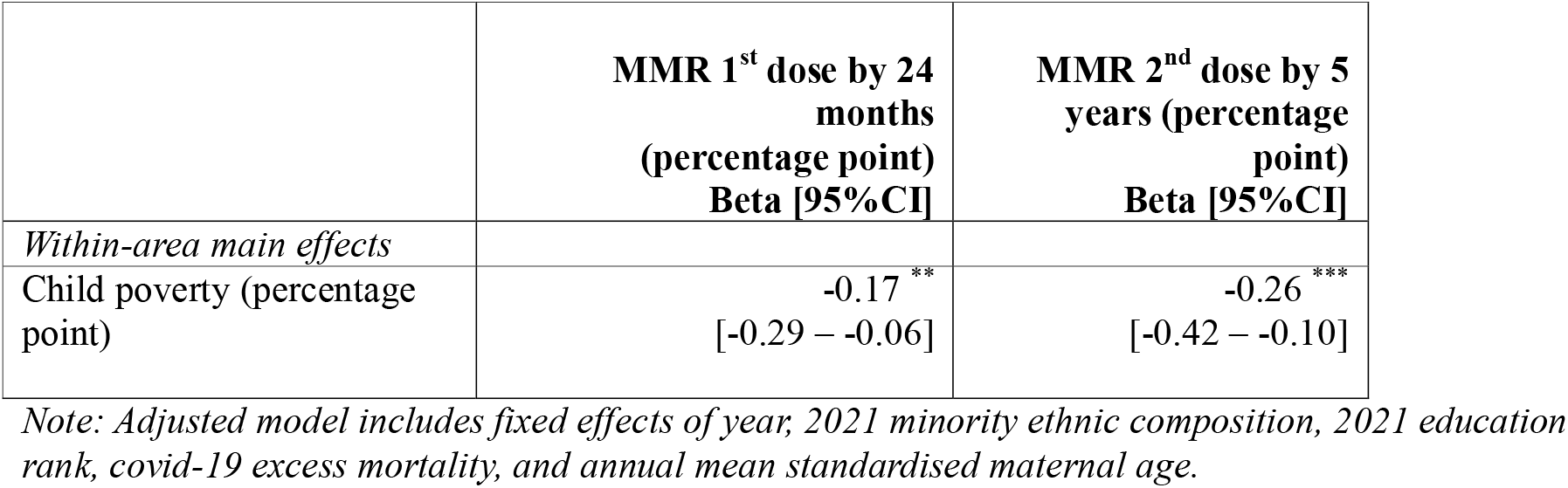
Adjusted estimates of the within-area association of child poverty on MMR vaccination rates.

|  | MMR 1<sup>st</sup> dose by 24 months<br>(percentage point)<br>Beta [95%CI] | MMR 2<sup>nd</sup> dose by 5 years (percentage point)<br>Beta [95%CI] |
| --- | --- | --- |
| *Within-area main effects* |  |  |
| Child poverty (percentage point) | -0.17 **<br>[-0.29 – -0.06] | -0.26 ***<br>[-0.42 – -0.10] |

Note: Adjusted model includes fixed effects of year, 2021 minority ethnic composition, 2021 education rank, covid-19 excess mortality, and annual mean standardised maternal age.

During the period of 2015 to 2024, there were 150143 more 5-year-old children who did not receive the MMR 2^nd^ dose, compared to the number of children had vaccination coverage remained at 2015 levels. The rise in child poverty contributed to 42705 (28.4%) of this preventable increase in children who did not receive the complete course of MMR vaccination.

### Between-area association of differences in child poverty on MMR rates

The interaction between the covid-19 and between-area child poverty term (MMR1 24M: χ2(1)=42.5, p<0.001; MMR2 5Y: χ2 (1)=44.8, p<0.001) indicates that areas with higher child poverty tended to have worse MMR rates after the pandemic, but a reverse association was seen before the pandemic (Beta MMR1 24M=-0.10 [0.12 – −0.07], MMR2 5Y= −0.13 [-0.17 – −0.09], p<0.001, see Table S4 and S5). Simple slopes of between-area associations of child poverty on MMR rates were small, i.e., a 1% difference in poverty was associated with less than 0.1% difference in MMR rates, and 95% confidence intervals overlapped with zero in both pre- and post-pandemic periods (Table S6). Adjusted and unadjusted parameter estimates of between-area child poverty were similar.

### Sensitivity analyses

In models lagging child poverty by a year (Table S7), confidence intervals of within- and between-area child poverty parameter estimates fell within the confidence intervals estimated in main analyses, indicating robustness of the models to temporal lags in measurement of the exposure and outcome. In fixed effect models, confidence intervals for within-area poverty in pre-covid19 (MMR1 24M: −0.18 [-0.35; −0.02]; −0.24[-0.48; −0.01] and post-covid19 (MMR1 24M −0.22[-0.35; −0.09]; MMR2 5Y - 0.32 [-0.47; −0.17]) periods were similar to the results of main models (Table S8 and S9). Similarly, including interactions did not meaningfully change the parameter estimates in main analyses for either within- or between-area associations of child poverty at the global mean (i.e., for average levels of exposure and included covariate) (Table S10). Interaction models suggest that vaccination rates may be more vulnerable to a rise in child poverty within the area in local authorities with a higher percentage of ethnic minorities (child poverty x ethnicity Beta MMR1 24M = −0.008 [-0.010; −0.006]; MMR2 5Y= −0.012 [-0.015; −0.009], p<0.001) or higher educational qualifications (child poverty x education for MMR2 5Y (Beta= −0.004 [-0.006; −0.002], p<0.001) (Table S10).

## Discussion

### Principal findings

On average between 2015 and 2024, coverage of the first dose of measles, mumps and rubella (MMR) vaccine at 24 months and the second dose at 5 years declined across English local authorities by 4 to 5 percentage points, and local authority child poverty increased by 5.6%. Increases in child poverty were associated with reductions in MMR uptake at both time points, with a stronger association observed for the second dose at 5 years. Over this 10-year period, the cumulative increase in children who did not receive the full MMR vaccination course was around 150,000, of which 28.4% could have been prevented, had child poverty remained at 2015 levels.

### Findings in context

A pre-covid-19 study in Italy found that cuts in public health expenditure were linked to declining MMR rates (24). Evidence from low-middle income countries also show that conditional cash transfer programs can temporarily boost vaccine uptake (25). Despite this, the authors could identify no research that has assessed the relationship between child poverty and childhood vaccine uptake. Here we provide that evidence, along with the extent to which a modifiable social determinant of health – child poverty - negatively impacts MMR vaccine uptake rates (24).

The relationship between child poverty and inequalities in vaccination rates has been recognised (26), but the pathways are complex. Increasing numbers of children into poverty are likely to worsen vaccination uptake through a multitude of individual, contextual and systems-level pathways (Figure S2). Inequalities to uptake are most pronounced, especially since the covid-19 pandemic, in socioeconomically deprived families, particularly children with young mothers aged <20 years, living in deprived areas, or Black or Black British households (27,28). Health literacy in part influences decisions and behaviours towards vaccination, further complicated by misinformation and lack of trust in both the government and health professionals, especially in underserved communities (29,30). However, health behaviours are constrained by contextual barriers including high childcare costs or unpaid time off work, insecure housing, along with various competing financial and social pressures – factors which often contribute to missed immunisations (26,31). Closures and funding cuts to public and child services contributing to vaccine literacy, access and uptake (e.g., health visiting, Sure Start), which impacted the most deprived areas the can further hinder the ability of health professionals to effectively support the needs of families in poverty (24,32).

### Implications for policy and practice

Despite a national immunisation collaboration between UKHSA, the Department of Health and Social Care, and NHS England, local service during the study period, delivery was fragmented, with organisations facing competing priorities and reduced budgets. The UKHSA Immunisation Equity Strategy for 2025-2030 and the NHS Vaccine Strategy, aim to enhance place-based community engagement, reduced fragmentation, and improved healthcare practitioner support (33), such as through partnerships between local NHS integrated care boards working in partnership with local authorities, community-led initiatives and trusted messengers (34). These strategies are expected to combat vaccine misinformation and improve uptake in disadvantaged groups. The 2026 MMR 2^nd^ dose schedule change, from the 3 years 4 months to 18-months is also expected to increase MMR 2^nd^ dose uptake, but it’s impact on inequalities remains uncertain (35,36).

While these strategies are welcome, they largely overlook the broader economic drivers of vaccination access such as child poverty and other social determinants of health. Economic analysis of policies designed to alleviate child poverty, should, in turn, assess both the financial impacts and downstream effects on healthcare services.

Applying the estimates from our findings to a stochastic transmission model (Supplemental Appendix S.3), we illustrate potential effects of poverty-driven declines in vaccine uptake on measles outbreak and health systems. A one percentage point increase in child poverty rates is linked to a 0.26 percentage point decline in MMR 2^nd^ dose uptake by age five – this would raise the effective reproduction number (R□) by approximately 1% and increase measles outbreak size by 6–14%. Since the UKHSA reported 2,911 confirmed measles cases in England in 2024 (4), a mid-range 10% increase in outbreak size due to increases in poverty would lead to roughly 291 (range: 175 to 408) additional confirmed cases. This is expected to cost £1.84 million (range: £1.12 million to £2.58 million) in direct and hospital costs, using economic modelling estimates from the previous UK measles outbreak (5). Broader impacts are seen in measles notifications, where a similar 10% increase would result in 1,116 additional notifications, costing around £2.71 million (range: £1.63 million to £3.80 million). These figures exclude additional costs such as productivity losses, outbreak-response spending, long-term complications, and mortality, indicating the true economic impact is significantly higher.

Overall, these findings suggest that addressing child poverty should be a central, preventative, focus of vaccination policies, likely to yield measurable public health and economic benefits by preventing outbreaks and reducing pressures on health systems. Economic policies that addressing poverty directly are needed, such as the recent removal of the two-child limit on Universal Credit and expanding access to benefits. Other approaches could be to target the systemic drivers of vaccine inequality associated with child poverty, including addressing barriers to access to health services and health information (Figure S2). For example, providing integrated early years services, restoring health visiting capacity are likely to be important targets for reducing fragmentation of vaccination services and other essential services for children facing poverty. While early years re-investment is progressing, funding for Family Hubs is still a fraction of what their predecessor, Sure Start Centres, received (8).

### Strengths and limitations

This is the first study to investigate the relationship between child poverty and vaccine uptake in England, using publicly available local authority data spanning a decade, and include improved official data on child poverty. Within-area analyses controlled for time-invariant local differences, economic trends, covid-19 impacts, and changes in maternal age. Area-level estimates are directly useful given the recent policy focus on place-based approaches for addressing health inequalities and vaccine uptake. However, the findings, especially between-area estimates, remain vulnerable to residual unmeasured confounding and as an ecological study, may not be generalisable to individual-level associations. To mitigate these issues, we adjusted for key confounders, including ethnic minority rates, since ethnic minority groups can have lower MMR vaccine uptake. Interaction effects revealed in sensitivity analyses suggest that individual-level data is needed to clarify the intersecting effects of child poverty with other influencing demographic factors.

## Conclusions

Rising child poverty is associated with reductions in childhood vaccine uptake. Since child poverty is largely driven by government economic policies it represents a modifiable factor for improving vaccination rates. These findings underscore the importance of an equity focused approach to vaccination, addressing the wider social determinants of vaccination uptake and strengthening healthcare delivery for vulnerable local communities.

## Ethical approval

Not required as data used for this study are anonymised, aggregated, and publicly available.

## Data availability statement

All data are open access and available through original sources. Vaccination data is available from NHS Digital (https://digital.nhs.uk/data-and-information/publications/statistical/nhs-immunisation-statisticsnhs) and data on child poverty from the Department for Work and Pensions (https://www.gov.uk/government/collections/children-in-low-income-families-local-area-statistics). R code and data for analysis are available on the Open Science Framework (DOI: https://osf.io/bd3fw/overview?view_only=37282a876b2c4650b938b06b6d7cedaa)

## Contribution

YWC is lead author and guarantor. Conceptualisation: YWC, DH, DTR; Methodology: YWC, LM, DH, DTR; Formal analysis: YWC; Data Curation: YWC; Writing – Original Draft: YWC, LM, OP, DH; Writing – Review & Editing: YWC, LM, OP, HS, MT, NF, MA, DH, DTR; Funding acquisition: YWC, DTR.

## References

1. Flatt A, Vivancos R, French N, Quinn S, Ashton M, Decraene V, et al. Inequalities in uptake of childhood vaccination in England, 2019-23: longitudinal study. BMJ. 2024 Dec 11;387:e079550. doi:10.1136/bmj-2024-079550

2. UK Health Security Agency. Measles: Historic confirmed cases, notifications and deaths [Internet]. 2025 [cited 2025 Dec 29]. Report No. Available from: https://www.gov.uk/government/publications/measles-historic-confirmed-cases-notifications-and-deaths/measles-historic-confirmed-cases-notifications-and-deaths

3. UK Health Security Agency. Why have we seen an increase in measles cases? 2019.

4. UK Health Security Agency. Infectious diseases impacting England: 2025 report [Internet]. 2025 [cited 2025 Dec 29]. Report No. Available from: https://www.gov.uk/government/publications/infectious-diseases-impacting-england-2025-report

5. Ghebrehewet S, Thorrington D, Farmer S, Kearney J, Blissett D, McLeod H, et al. The economic cost of measles: Healthcare, public health and societal costs of the 2012–13 outbreak in Merseyside, UK. Vaccine. 2016 Apr;34(15):1823–31. doi:10.1016/j.vaccine.2016.02.029

6. Morton A. The role of the health visitor: where are we now? Paediatrics and Child Health. 2024 Jul 1;34(7):234–8. doi:10.1016/j.paed.2024.04.006

7. Institute of Health Visiting. From disparity to opportunitiy: The case for rebuilding health visiting [State of Health Visiting, UK survey report]. 2025 Jan. Report No. doi:https://ihv.org.uk/wp-content/uploads/2025/01/State_of_Health_Visiting_Report_2024_FINAL_VERSION_22.01.25_compressed.pdf

8. Carneiro P, Cattan S, Ridpath N. The short- and medium-term impacts of Sure Start on educational outcomes [Internet]. 2024 Apr [cited 2024 Nov 26]. Report No. Available from: https://ifs.org.uk/publications/short-and-medium-term-impacts-sure-start-educational-outcomes

9. Hourston P. Cost of living crisis [Explainer] [Internet]. Institute for Government; 2022 Feb [cited 2026 Feb 19]. Report No. Available from: https://www.instituteforgovernment.org.uk/explainer/cost-living-crisis

10. Bennett DL, Schlüter DK, Melis G, Bywaters P, Alexiou A, Barr B, et al. Child poverty and children entering care in England, 2015–20: a longitudinal ecological study at the local area level. The Lancet Public Health. 2022 Jun;7(6):e496–503. doi:10.1016/S2468-2667(22)00065-2

11. Taylor-Robinson D, Lai ETC, Wickham S, Rose T, Norman P, Bambra C, et al. Assessing the impact of rising child poverty on the unprecedented rise in infant mortality in England, 2000–2017: time trend analysis. BMJ Open. 2019 Oct;9(10):e029424. doi:10.1136/bmjopen-2019-029424

12. Child Poverty Action Group. CPAG’s response to the latest poverty statistics [Internet]. 2025. Report No. Available from: https://cpag.org.uk/sites/default/files/2025-03/Child_poverty_statistics_2025.pdf

13. Child Poverty Action Group. Reducing child poverty: role of the two-child limit [Internet]. 2025. Report No. Available from: https://cpag.org.uk/sites/default/files/2025-04/Reducing_child_poverty_role_of_two-child_limit.pdf

14. Office for National Statistics. Area type definitions Census 2021. Report No.

15. NHS England. Childhood Vaccination Coverage Statistics [Internet]. NHS Digital; 2024. Available from: https://digital.nhs.uk/data-and-information/publications/statistical/nhs-immunisation-statisticsnhs

16. Department for Work and Pensions. atChildren in low income families: local area statistics [Internet]. 2024 [cited 2024 Nov 14]. Available from: https://www.gov.uk/government/collections/children-in-low-income-families-local-area-statistics

17. Office for National Statistics. Population estimates - local authority based by single year of age - Nomis - Official Census and Labour Market Statistics [Internet]. Nomis - Official Census and Labour Market Statistics; 2023 [cited 2024 Nov 14]. Available from: https://www.nomisweb.co.uk/datasets/pestsyoala

18. Wooldridge. Econometric Analysis of Cross Section and Panel Data [Internet]. Cambridge, Massachusetts; London, England: The MIT Press; 2001. 276–285 p. Available from: https://mitpress.mit.edu/9780262232586/econometric-analysis-of-cross-section-and-panel-data/

19. Department of Health and Social CAre, Office for National Statistics. Direct and indirect health impacts of COVID-19 in England: emerging Omicron impacts [Internet]. 2022. Report No. Available from: https://www.gov.uk/government/publications/direct-and-indirect-health-impacts-of-covid-19-in-england-emerging-omicron-impacts

20. Office for Health Improvement and Disparities. Excess mortality in England and English regions: March 2020 to December 2023 [Internet]. 2024 [cited 2024 Nov 14]. Available from: https://www.gov.uk/government/statistics/excess-mortality-in-england-and-english-regions

21. Office for National Statistics. Ethnic group, England and Wales: Census 2021 [Internet]. 2021 [cited 2024 Nov 27]. Report No. Available from: https://www.ons.gov.uk/peoplepopulationandcommunity/culturalidentity/ethnicity/bulletins/ethnicgroupenglandandwales/census2021

22. Office for National Statistics. Live births in England and Wales: birth rates down to local authority areas [Internet]. Nomis - Official Census and Labour Market Statisticspop; 2024. Available from: https://www.nomisweb.co.uk/datasets/lebirthrates

23. Schunck R. Within and between Estimates in Random-Effects Models: Advantages and Drawbacks of Correlated Random Effects and Hybrid Models. The Stata Journal. 2013 Mar 1;13(1):65–76. doi:10.1177/1536867X1301300105

24. Toffolutti V, McKee M, Melegaro A, Ricciardi W, Stuckler D. Austerity, measles and mandatory vaccination: cross-regional analysis of vaccination in Italy 2000–14. European Journal of Public Health. 2019 Feb 1;29(1):123–7. doi:10.1093/eurpub/cky178

25. Saunders MJ, Pereboom M, Alvarez JL, Sherlock M, Gadroen K. Incentives in immunisation campaigns in low- and middle-income countries: a scoping review mapping evidence on effectiveness and unintended consequences. BMJ Glob Health. 2025 Jun;10(6):e019662. doi:10.1136/bmjgh-2025-019662

26. Department of Health and Social Care. Chief Medical Officer’s annual report 2025: infections [Internet]. 2025 [cited 2025 Dec 29]. Report No. Available from: https://www.gov.uk/government/publications/chief-medical-officers-annual-report-2025-infections

27. Skirrow H, Foley K, Costelloe C, Bedford H, Lewis C, Whittaker E, et al. Maternal predictors of timeliness & uptake of Measles, Mumps & Rubella vaccine: A birth cohort study. Eur J Public Health. 2023 Oct 24;33(Suppl 2):ckad160.139. doi:10.1093/eurpub/ckad160.139 PubMed PMID: null; PubMed Central PMCID: PMC10596084.

28. Harris C, Bird C, Saavedra-Campos M, Chatt C, Booth E, Proto W, et al. Impact of a measles outbreak on a UK children’s emergency department and the public health response: a retrospective observational study. Journal of Hospital Infection. 2025 Nov;165:41–7. doi:10.1016/j.jhin.2025.07.020

29. Adhikari B, Yeong Cheah P, von Seidlein L. Trust is the common denominator for COVID-19 vaccine acceptance: A literature review. Vaccine: X. 2022 Dec 1;12:100213. doi:10.1016/j.jvacx.2022.100213

30. Dubé E, Gagnon D, MacDonald NE. Strategies intended to address vaccine hesitancy: Review of published reviews. Vaccine. 2015 Aug 14; WHO Recommendations Regarding Vaccine Hesitancy 33(34):4191–203. doi:10.1016/j.vaccine.2015.04.041

31. Royal College of Paediatrics and Child Health (RCPCH). Vaccination in the UK: Access, uptake and equity. 2025. Report No.

32. Finch D, Gazzillo A, Vriend M. Investing in the public health grant - The Health Foundation [Internet]. 2025 [cited 2026 Mar 2]. Report No. Available from: https://www.health.org.uk/reports-and-analysis/analysis/investing-in-the-public-health-grant

33. NHS England. NHS England□» NHS vaccination strategy [Internet]. [cited 2026 Feb 19]. Report No. Available from: https://www.england.nhs.uk/long-read/nhs-vaccination-strategy/

34. Essa-Hadad J, Gorelik Y, Vervoort J, Jansen D, Edelstein M. Improving childhood vaccination among minority populations in middle- and high-income countries: a realist review of health system interventions. J Epidemiol Community Health. 2026 Jan 23;jech-2025-225099. doi:10.1136/jech-2025-225099

35. UK Health Security Agency. Changes to the childhood vaccination schedule from January 2026 [Internet]. 2025 Dec 30 [cited 2026 Feb 19]. Available from: https://ukhsa.blog.gov.uk/2025/12/30/changes-to-the-childhood-vaccination-schedule-from-january-2026/

36. Lacy J, Tessier E, Andrews N, White J, Ramsay M, Edelstein M. Impact of an accelerated measles-mumps-rubella (MMR) vaccine schedule on vaccine coverage: An ecological study among London children, 2012–2018. Vaccine. 2022 Jan 24;40(3):444–9. doi:10.1016/j.vaccine.2021.12.011

