## Supplemental Appendices for "Child poverty and declining measles, mumps and rubella (MMR) vaccination in England, 2015 to 2024. A longitudinal ecological study at local area level"

**Supplemental materials**

**S.1. Supplemental methods**

Covariates were scaled to capture a 1 percentage-point, 1 percentile, 1 year change, or 1 standard deviation change in ethnic minority percentage, education qualification, maternal age or excess mortality respectively. Covariates were centred so parameter estimates are estimated at the mean (over area and time) of child poverty and maternal age, the mean (over area) of ethnicity and excess mortality, and 50^th^ percentile of education rank.

Three types of interactions are possible in within-between models: interactions of between-area child poverty and each covariate (between-between interactions), within-area child poverty and each time-invariant covariate (within-between interactions) and within-area child poverty and time-varying covariate (within-within interaction of child poverty and maternal age).

***Between-between interactions of covariate and child poverty*** capture how covariate levels varying between LA influence **between-area** associations of child poverty on the outcome. For example, at higher ethnic minority populations, the association between child poverty on the outcome might be stronger.

***Between-within interactions of covariate and child poverty*** capture how covariate levels varying between LA influence **within-area** associations of child poverty on the outcome. For example, in areas with higher ethnic minority population, changes within-LA in child poverty on within-LA changes in the outcome might be stronger.

***Within-within interactions of time-varying maternal age and child poverty*** capture how **within-area** changes in maternal age may influence how strongly **within-area** changes in child poverty influence the outcome. For example, if child poverty and maternal age in an LA simultaneously increase, whether we see a larger increase in MMR rates compared to if only child poverty increases. The between-between and within-within interaction terms for time-varying maternal age and child poverty were generated using the double demeaning approach (Gisselmann & Schmidt-Catran, 2022).

We do not focus on interactions in the main analysis because most covariates were available at a single time point and the area-level nature of the data means the findings are prone to ecological fallacy. Inclusion of interaction effects were considered using a step-down approach considering model fit and statistical significance of parameter estimates. Within-between interactions of child poverty x ethnicity, child poverty x education, and between-between interaction of child poverty x ethnicity were statistically significant. Removal of all other non-statistically significant interactions with child poverty did not worsen model fit (Table S1). We visualised interactions by plotting the simple slopes of within-area or between-area child poverty at quintiles (see Table S2) of the interacting variable*.*  Between-area interactions of child poverty and ethnicity were included for completeness, but are not discussed further in the main results as between-area estimates are less robust, and interactions may be driven by trends in the smaller number of local authorities at the intersections of high or low child poverty and ethnic minorities.

**References for Supplemental Materials S.1.**

Giesselmann M, Schmidt-Catran AW. Interactions in Fixed Effects Regression Models. Sociological Methods & Research. 2022 Aug 1;51(3):1100–27.

**S.2. Supplemental tables and figures**

**Table S1. Model fit indices for models considering interactions in a step-down approach**

| **Outcome** | **Model** | **df** | **AIC** | **BIC** | **Log Likelihood** | **χ^2^** | **p** |
| --- | --- | --- | --- | --- | --- | --- | --- |
| MMR 24M | All interactions |  |  |  |  |  |  |
|  | minus mortality x poverty interactions | 28 | 6750.846 | 6899.145 | -3347.42 | -9.97 | >1.000 |
|  | minus age x poverty interactions | 26 | 6736.877 | 6874.584 | -3342.44 | -6.61 | >1.000 |
|  | minus education x mean_pov interaction | 24 | 6726.264 | 6853.378 | -3339.13 | -12.52 | >1.000 |
| MMR 5Y | All interactions | 23 | 6711.749 | 6833.566 | -3332.87 |  |  |
|  | minus mortality x poverty interactions | 28 | 7631.837 | 7780.136 | -3787.92 | -6.06 | >1.000 |
|  | minus age x poverty interactions | 26 | 7621.779 | 7759.485 | -3784.89 | -4.78 | >1.000 |
|  | minus education x mean_pov interaction | 24 | 7613.001 | 7740.115 | -3782.5 | -11.51 | >1.000 |
|  |  | 23 | 7599.486 | 7721.303 | -3776.74 |  |  |

**Table S2. Quintiles of child poverty, ethnic minority percent and education rank**

| **Quintile** | **Child poverty (%)** | **Ethnic Minority (%)** | **Education rank** |
| --- | --- | --- | --- |
| 1 | 2.7 | 6.8 | 16 |
| 2 | 13.0 | 11.5 | 46 |
| 3 | 17.1 | 21.3 | 75 |
| 4 | 22.5 | 34.4 | 104 |
| 5 | 30.2 | 64.7 | 133 |

**Table S3. Area-level mean and 95% confidence intervals of child poverty, MMR 1^st^ dose by 24 months and MMR 2^nd^ dose by 5 years**

| **Year** | **Child poverty (%) mean [95%CI]** | **MMR 1^st^ dose by 24 months (%) mean [95%CI]** | **MMR 2^nd^ dose by 5 years (%) mean [95%CI]** |
| --- | --- | --- | --- |
| 2015 | 14.7 [13.4;15.9] | 92.4 [91.8;93.1] | 88.8 [87.9;89.8] |
| 2016 | 15.7 [14.4;17.1] | 91.9 [91.2;92.6] | 88.5 [87.4;89.5] |
| 2017 | 15.6 [14.3;17] | 91.6 [90.7;92.4] | 87.6 [86.5;88.7] |
| 2018 | 17.8 [16.3;19.4] | 91.1 [90.4;91.8] | 86.9 [85.9;88] |
| 2019 | 16.7 [15.2;18.1] | 90.3 [89.5;91.1] | 86.2 [85.1;87.3] |
| 2020 | 19.5 [17.8;21.1] | 90.6 [89.9;91.4] | 86.5 [85.4;87.7] |
| 2021 | 17.2 [15.7;18.6] | 90.3 [89.5;91.1] | 86.3 [85.1;87.6] |
| 2022 | 17.7 [16.1;19.2] | 89.1 [88.1;90.1] | 85.5 [84.3;86.7] |
| 2023 | 19.6 [17.8;21.3] | 89.2 [88.4;90.1] | 84.3 [83.1;85.5] |
| 2024 | 20.3 [18.5;22.0] | 88.8 [88;89.7] | 83.6 [82.4;84.8] |

**Table S4. Unadjusted and adjusted model coefficients of child poverty on MMR 1^st^ dose by 24 months**

|  | **MMR 1^st^ dose by 24 months**  **Unadjusted model** | | | **MMR 1^st^ dose by 24 months**  **Adjusted model** | | |
| --- | --- | --- | --- | --- | --- | --- |
| *Predictors* | *Estimates* | *CI* | *p* | *Estimates* | *CI* | *p* |
| ***Within-area associations*** |  |  |  |  |  |  |
| Child poverty (%) | -0.18 ^**^ | -0.29 – -0.06 | **0.003** | -0.17 ^**^ | -0.29 – -0.06 | **0.003** |
| Maternal age (years) |  |  |  | -0.37 | -1.09 – 0.35 | 0.311 |
| ***Between-area associations*** |  |  |  |  |  |  |
| Child poverty (%) | 0.04 | -0.04 – 0.12 | 0.351 | 0.04 | -0.02 – 0.10 | 0.227 |
| Child poverty × post-covid19 [Ref: pre-covid-19] | -0.10 ^***^ | -0.12 – -0.07 | **<0.001** | -0.10 ^***^ | -0.12 – -0.07 | **<0.001** |
| Minority ethnicity (%) |  |  |  | -0.14 ^***^ | -0.18 – -0.11 | **<0.001** |
| Educational qualification (percentile) |  |  |  | 0.00 | -0.02 – 0.02 | 0.849 |
| Maternal age (years) |  |  |  | -0.77 ^*^ | -1.48 – -0.07 | **0.032** |
| covid-19 excess mortality |  |  |  | 0.03 | -0.41 – 0.46 | 0.899 |
| ***Fixed effects of time period*** |  |  |  |  |  |  |
| 2015 [Ref: 2019] | -0.32 | -0.80 – 0.15 | 0.183 | -0.28 | -0.77 – 0.20 | 0.253 |
| 2016 [Ref: 2019] | -0.68 ^**^ | -1.16 – -0.21 | **0.005** | -0.62 ^*^ | -1.11 – -0.13 | **0.014** |
| 2017 [Ref: 2019] | -0.71 ^*^ | -1.29 – -0.12 | **0.018** | -0.61 ^*^ | -1.23 – -0.00 | **0.05** |
| 2018 [Ref: 2019] | -1.78 ^***^ | -2.30 – -1.27 | **<0.001** | -1.66 ^***^ | -2.23 – -1.09 | **<0.001** |
| 2020 [Ref: 2019] | 0.71 ^*^ | 0.07 – 1.36 | **0.029** | 0.86 ^*^ | 0.16 – 1.56 | **0.016** |
| 2021 [Ref: 2019] | 0 | -0.60 – 0.60 | 0.998 | 0.22 | -0.51 – 0.96 | 0.556 |
| 2022 [Ref: 2019] | -1.10 ^***^ | -1.70 – -0.50 | **<0.001** | -0.89 ^*^ | -1.61 – -0.17 | **0.015** |
| 2023 [Ref: 2019] | -0.63 | -1.27 – 0.02 | 0.058 | -0.41 | -1.18 – 0.36 | 0.298 |
| 2024 [Ref: 2019] | -0.92 ^**^ | -1.61 – -0.24 | **0.008** | -0.68 | -1.51 – 0.15 | 0.108 |
| (Intercept) | 91.27 ^***^ | 89.67 – 92.86 | **<0.001** | 114.79 ^***^ | 92.45 – 137.13 | **<0.001** |
| **Random Effects** |  |  |  |  |  |  |
| σ^2^ | 4.06 |  |  | 4.06 |  |  |
| τ_00_ | 19.90 _AreaName_ |  |  | 5.22 _AreaName_ |  |  |
| ICC | 0.83 |  |  | 0.56 |  |  |
| N | 148 _AreaName_ |  |  | 148 _AreaName_ |  |  |
| Observations | 1475 |  |  | 1475 |  |  |
| Marginal R^2^ / Conditional R^2^ | 0.068 / 0.842 | |  | 0.640 / 0.842 | |  |
| AIC | 6890.276 |  |  | 6729.209 |  |  |
| ** p<0.05   ** p<0.01   *** p<0.001* |  |  |  |  |  |  |

**Table S5. Unadjusted and adjusted model coefficients of child poverty on MMR 2^nd^ dose by 5 years**

|  | **MMR 2^nd^ dose by 5 years**  **Unadjusted model** | | | **MMR 2^nd^ dose by 5 years**  **Adjusted model** | | |
| --- | --- | --- | --- | --- | --- | --- |
| *Predictors* | *Estimates* | *CI* | *p* | *Estimates* | *CI* | *p* |
| ***Within-area associations*** |  |  |  |  |  |  |
| Child poverty (%) | -0.27 ^***^ | -0.43 – -0.12 | **0.001** | -0.26 ^**^ | -0.42 – -0.10 | **0.001** |
| Maternal age (years) |  |  |  | -1.44 ^**^ | -2.41 – -0.47 | **0.004** |
| ***Between-area associations*** |  |  |  |  |  |  |
| Child poverty (%) | 0.08 | -0.04 – 0.19 | 0.184 | 0.09 | -0.01 – 0.18 | 0.065 |
| Child poverty × post-covid19 [Ref: pre-covid-19] | -0.13 ^***^ | -0.17 – -0.09 | **<0.001** | -0.13 ^***^ | -0.17 – -0.10 | **<0.001** |
| Minority ethnicity (%) |  |  |  | -0.21 ^***^ | -0.26 – -0.16 | **<0.001** |
| Educational qualification (percentile) |  |  |  | -0.01 | -0.04 – 0.02 | 0.588 |
| Maternal age (years) |  |  |  | -0.87 | -1.93 – 0.20 | 0.109 |
| covid-19 excess mortality |  |  |  | 0.12 | -0.54 – 0.77 | 0.728 |
| ***Fixed effects of time period*** |  |  |  |  |  |  |
| 2015 [Ref: 2019] | -0.06 | -0.71 – 0.59 | 0.855 | 0.1 | -0.55 – 0.76 | 0.756 |
| 2016 [Ref: 2019] | -0.94 ^**^ | -1.58 – -0.30 | **0.004** | -0.69 ^*^ | -1.35 – -0.03 | **0.042** |
| 2017 [Ref: 2019] | -1.04 ^*^ | -1.84 – -0.25 | **0.01** | -0.69 | -1.51 – 0.14 | 0.105 |
| 2018 [Ref: 2019] | -2.06 ^***^ | -2.76 – -1.36 | **<0.001** | -1.59 ^***^ | -2.35 – -0.82 | **<0.001** |
| 2020 [Ref: 2019] | 1.33 ^**^ | 0.47 – 2.20 | **0.003** | 1.90 ^***^ | 0.95 – 2.84 | **<0.001** |
| 2021 [Ref: 2019] | 0.5 | -0.31 – 1.31 | 0.227 | 1.36 ^**^ | 0.36 – 2.35 | **0.007** |
| 2022 [Ref: 2019] | -0.19 | -1.00 – 0.63 | 0.653 | 0.61 | -0.36 – 1.58 | 0.217 |
| 2023 [Ref: 2019] | -0.90 ^*^ | -1.78 – -0.03 | **0.044** | -0.06 | -1.10 – 0.98 | 0.913 |
| 2024 [Ref: 2019] | -1.38 ^**^ | -2.30 – -0.45 | **0.004** | -0.42 | -1.54 – 0.70 | 0.461 |
| (Intercept) | 86.75 ^***^ | 84.48 – 89.02 | **<0.001** | 112.54 ^***^ | 78.91 – 146.17 | **<0.001** |
| **Random Effects** |  |  |  |  |  |  |
| σ^2^ | 7.46 |  |  | 7.41 |  |  |
| τ_00_ | 40.62 _AreaName_ |  |  | 12.01 _AreaName_ |  |  |
| ICC | 0.84 |  |  | 0.62 |  |  |
| N | 148 _AreaName_ |  |  | 148 _AreaName_ |  |  |
| Observations | 1475 |  |  | 1475 |  |  |
| Marginal R^2^ / Conditional R^2^ | 0.064 / 0.855 | |  | 0.640 / 0.842 | |  |
| AIC | 7795.096 |  |  | 7638.216 |  |  |
| ** p<0.05   ** p<0.01   *** p<0.001* |  |  |  |  |  |  |

**Table S6. Simple slopes of between-area child poverty in 2015-2019 (pre-covid19) and 2020 – 2024 (post-covid19)**

|  | **covid19 period** | **Simple slope of between-area child poverty**  **Estimate[95% CI]** | **SE** | **df** | **t** | **p** |
| --- | --- | --- | --- | --- | --- | --- |
| **MMR 1^st^ dose by 24 months** | 2015 - 2019 | 0.04 [-0.03 – 0.10] | 0.03 | 171.75 | 1.19 | 0.236 |
|  | 2020 - 2024 | -0.06 [-0.12 – 0.01] | 0.03 | 171.71 | -1.78 | 0.077 |
| **MMR 2^nd^ dose by 5 years** | 2015 - 2019 | 0.09 [-0.01 – 0.18] | 0.05 | 168.30 | 1.82 | 0.071 |
|  | 2020 - 2024 | -0.05 [-0.14 – 0.05] | 0.05 | 168.27 | -0.95 | 0.342 |

**Table S7. Sensitivity analyses model coefficients of lagged child poverty on MMR 1^st^ dose by 24 months and MMR 2^nd^ dose by 5 years**

|  | **MMR 1^st^ dose by 24 months  Adjusted model with lagged child poverty** | | | **MMR 2^nd^ dose by 5 years**  **Adjusted model with lagged child poverty** | | |
| --- | --- | --- | --- | --- | --- | --- |
| *Predictors* | *Estimates* | *CI* | *p* | *Estimates* | *CI* | *p* |
| ***Within-area associations*** |  |  |  |  |  |  |
| Child poverty (%) | -0.13 ^*^ | -0.25 – -0.01 | **0.031** | -0.12 | -0.28 – 0.04 | 0.153 |
| Maternal age (years) | -0.19 | -0.96 – 0.58 | 0.621 | -1.04 ^*^ | -2.08 – -0.01 | **0.049** |
| ***Between-area associations*** |  |  |  |  |  |  |
| Child poverty (%) | 0.03 | -0.04 – 0.09 | 0.439 | 0.07 | -0.02 – 0.17 | 0.129 |
| Child poverty × post-covid19 [Ref: pre-covid-19] | -0.10 ^***^ | -0.13 – -0.07 | **<0.001** | -0.16 ^***^ | -0.19 – -0.12 | **<0.001** |
| Minority ethnicity (%) | -0.15 ^***^ | -0.18 – -0.11 | **<0.001** | -0.21 ^***^ | -0.26 – -0.16 | **<0.001** |
| Educational qualification (percentile) | 0 | -0.02 – 0.02 | 0.83 | -0.01 | -0.04 – 0.02 | 0.538 |
| Maternal age (years) | -0.78 ^*^ | -1.51 – -0.05 | **0.036** | -0.89 | -1.98 – 0.20 | 0.11 |
| covid-19 excess mortality | 0.02 | -0.43 – 0.47 | 0.938 | 0.11 | -0.57 – 0.78 | 0.756 |
| ***Fixed effects of time period*** |  |  |  |  |  |  |
| 2016 [Ref: 2019] | -0.18 | -0.65 – 0.29 | 0.46 | -0.6 | -1.24 – 0.04 | 0.065 |
| 2017 [Ref: 2019] | -0.59 ^*^ | -1.07 – -0.11 | **0.016** | -1.25 ^***^ | -1.90 – -0.61 | **<0.001** |
| 2018 [Ref: 2019] | -1.15 ^***^ | -1.77 – -0.53 | **<0.001** | -1.60 ^***^ | -2.43 – -0.76 | **<0.001** |
| 2020 [Ref: 2019] | -0.97 ^***^ | -1.54 – -0.40 | **0.001** | -1.36 ^***^ | -2.13 – -0.59 | **0.001** |
| 2021 [Ref: 2019] | 0.89 ^*^ | 0.16 – 1.61 | **0.016** | 1.57 ^**^ | 0.59 – 2.54 | **0.002** |
| 2022 [Ref: 2019] | -0.57 | -1.31 – 0.18 | 0.138 | 0.66 | -0.35 – 1.67 | 0.197 |
| 2023 [Ref: 2019] | -0.38 | -1.11 – 0.36 | 0.315 | -0.55 | -1.54 – 0.43 | 0.271 |
| 2024 [Ref: 2019] | -0.54 | -1.34 – 0.26 | 0.189 | -0.95 | -2.03 – 0.14 | 0.086 |
| (Intercept) | 114.89 ^***^ | 91.77 – 138.00 | **<0.001** | 113.67 ^***^ | 79.05 – 148.29 | **<0.001** |
| **Random Effects** |  |  |  |  |  |  |
| σ^2^ | 3.78 |  |  | 6.87 |  |  |
| τ_00_ | 5.61 _AreaName_ |  |  | 12.75 _AreaName_ |  |  |
| ICC | 0.6 |  |  | 0.65 |  |  |
| N | 148 _AreaName_ |  |  | 148 _AreaName_ |  |  |
| Observations | 1328 |  |  | 1328 |  |  |
| Marginal R^2^ / Conditional R^2^ | 0.644 / 0.857 | |  | 0.640 / 0.842 | |  |
| AIC | 6014.871 |  |  | 6827.144 |  |  |
| ** p<0.05   ** p<0.01   *** p<0.001* |  |  |  |  |  |  |

**Table S8. Mundlak specification MMR 1^st^ dose by 24 months, stratified by pre- and post-covid19 period**

|  | **MMR 1^st^ dose by 24 months (2015 - 2019)** | | | **MMR 1^st^ dose by 24 months (2020 - 2024)** | | |
| --- | --- | --- | --- | --- | --- | --- |
| Predictors | *Estimates* | *CI* | *Estimates* | *CI* | *Estimates* | *CI* |
| ***Time-varying variables*** |  |  |  |  |  |  |
| Child poverty (%) | -0.18 | -0.35 – -0.02 | -0.18 | -0.35 – -0.02 | -0.18 | -0.35 – -0.02 |
| Maternal age (years) | 0.20 | -0.97 – 1.38 | 0.20 | -0.97 – 1.38 | 0.20 | -0.97 – 1.38 |
| ***Fixed effects of timeperiod*** |  |  |  |  |  |  |
| 2016 [Ref: 2015] | -0.34 | -0.81 – 0.13 | -0.34 | -0.81 – 0.13 | -0.34 | -0.81 – 0.13 |
| 2017 [Ref: 2015] | -0.71 | -1.21 – -0.22 | -0.71 | -1.21 – -0.22 | -0.71 | -1.21 – -0.22 |
| 2018 [Ref: 2015] | -0.74 | -1.48 – 0.00 | -0.74 | -1.48 – 0.00 | -0.74 | -1.48 – 0.00 |
| 2019 [Ref: 2015] | -1.84 | -2.51 – -1.17 | -1.84 | -2.51 – -1.17 | -1.84 | -2.51 – -1.17 |
| 2016 [Ref: 2020] |  |  |  |  |  |  |
| 2017 [Ref: 2020] |  |  |  |  |  |  |
| 2018 [Ref: 2020] |  |  |  |  |  |  |
| 2019 [Ref: 2020] |  |  |  |  |  |  |
| Observations | 735 |  | 735 |  | 735 |  |
| R2 | 0.173 |  | 0.173 |  | 0.173 |  |
| R2 Adj. | -0.043 |  | -0.043 |  | -0.043 |  |
| AIC | 2805.1 |  | 2805.1 |  | 2805.1 |  |

**Table S9. Mundlak specification MMR 2^nd^ dose by 5 years, stratified by pre- and post-covid19 period**

|  | **MMR 2^nd^ dose by 5 years (2015 - 2019)** | | | **MMR 2^nd^ dose by 5 years (2020 - 2024)** | | |
| --- | --- | --- | --- | --- | --- | --- |
| Predictors | *Estimates* | *CI* | *p* | *Estimates* | *CI* | *p* |
| ***Time-varying variables*** |  |  |  |  |  |  |
| Child poverty (%) | -0.24 | -0.48 – -0.01 | **0.044*** | -0.32 | -0.47 – -0.17 | **<0.001***** |
| Maternal age (years) | -1.19 | -2.89 – 0.51 | 0.171 | -0.04 | -1.02 – 0.94 | 0.934 |
| ***Fixed effects of timeperiod*** |  |  |  |  |  |  |
| 2016 [Ref: 2015] | 0.06 | -0.63 – 0.74 | 0.874 |  |  |  |
| 2017 [Ref: 2015] | -0.75 | -1.46 – -0.04 | 0.039 |  |  |  |
| 2018 [Ref: 2015] | -0.81 | -1.87 – 0.26 | 0.138 |  |  |  |
| 2019 [Ref: 2015] | -1.71 | -2.67 – -0.75 | <0.001 |  |  |  |
| 2016 [Ref: 2020] |  |  |  | -0.93 | -1.49 – -0.37 | 0.001 |
| 2017 [Ref: 2020] |  |  |  | -1.60 | -2.10 – -1.10 | <0.001 |
| 2018 [Ref: 2020] |  |  |  | -2.22 | -2.65 – -1.80 | <0.001 |
| 2019 [Ref: 2020] |  |  |  | -2.66 | -3.14 – -2.19 | <0.001 |
| Observations | 735 |  |  | 740 |  |  |
| R2 | 0.153 |  |  | 0.385 |  |  |
| R2 Adj. | -0.068 |  |  | 0.224 |  |  |
| AIC | 3345.5 |  |  | 2701.2 |  |  |

**Table S10. Sensitivity analyses model coefficients of child poverty on MMR 1^st^ dose by 24 months and MMR 2^nd^ dose by 5 years including interactions**

|  | **MMR 1^st^ dose by 24 months Adjusted model with interactions** | | | **MMR 2^nd^ dose by 5 years**  **Adjusted model with interactions** | | |
| --- | --- | --- | --- | --- | --- | --- |
| *Predictors* | *Estimates* | *CI* | *p* | *Estimates* | *CI* | *p* |
| ***Within-area associations*** |  |  |  |  |  |  |
| Child poverty (%) | -0.20 ^***^ | -0.32 – -0.09 | **<0.001** | -0.30 ^***^ | -0.45 – -0.15 | **<0.001** |
| Maternal age (years) | -0.06 | -0.77 – 0.65 | 0.861 | -0.54 | -1.53 – 0.44 | 0.28 |
| ***Between-area associations*** |  |  |  |  |  |  |
| Child poverty (%) | 0.03 | -0.03 – 0.09 | 0.288 | 0.08 | -0.01 – 0.17 | 0.073 |
| Child poverty × post-covid19 [Ref: pre-covid-19] | -0.091 ^***^ | -0.119 – -0.062 | **<0.001** | -0.133 ^***^ | -0.171 – -0.095 | **<0.001** |
| Minority ethnicity (%) | -0.24 ^***^ | -0.30 – -0.18 | **<0.001** | -0.34 ^***^ | -0.43 – -0.25 | **<0.001** |
| Educational qualification (percentile) | 0.01 | -0.01 – 0.03 | 0.55 | 0 | -0.03 – 0.03 | 0.845 |
| Maternal age (years) | -0.86 ^*^ | -1.53 – -0.19 | **0.012** | -0.99 | -2.01 – 0.03 | 0.057 |
| covid-19 excess mortality | -0.46 | -0.93 – 0.02 | 0.061 | -0.56 | -1.28 – 0.17 | 0.133 |
| ***Within-Between interactions*** |  |  |  |  |  |  |
| Child poverty × minority ethnicity | -0.008 ^***^ | -0.010 – -0.006 | **<0.001** | -0.012 ^***^ | -0.015 – -0.009 | **<0.001** |
| Child poverty × education |  |  |  | -0.004 ^***^ | -0.006 – -0.002 | **<0.001** |
| ***Between-Between interactions*** |  |  |  |  |  |  |
| Child poverty x minority ethnicity | 0.005 ^***^ | 0.003 – 0.008 | **<0.001** | 0.007 ^***^ | 0.003 – 0.012 | **<0.001** |
| ***Fixed effects of time period*** |  |  |  |  |  |  |
| 2015 [Ref: 2019] | -0.28 | -0.76 – 0.19 | 0.243 | 0.04 | -0.60 – 0.68 | 0.898 |
| 2016 [Ref: 2019] | -0.65 ^**^ | -1.13 – -0.17 | **0.009** | -0.82 ^*^ | -1.47 – -0.17 | **0.013** |
| 2017 [Ref: 2019] | -0.6 | -1.20 – 0.00 | 0.052 | -0.8 | -1.61 – 0.01 | 0.054 |
| 2018 [Ref: 2019] | -1.70 ^***^ | -2.26 – -1.15 | **<0.001** | -1.82 ^***^ | -2.57 – -1.06 | **<0.001** |
| 2020 [Ref: 2019] | 0.75 ^*^ | 0.06 – 1.44 | **0.032** | 1.64 ^***^ | 0.71 – 2.56 | **0.001** |
| 2021 [Ref: 2019] | 0.04 | -0.68 – 0.77 | 0.911 | 0.93 | -0.05 – 1.91 | 0.062 |
| 2022 [Ref: 2019] | -1.07 ^**^ | -1.78 – -0.36 | **0.003** | 0.2 | -0.75 – 1.16 | 0.674 |
| 2023 [Ref: 2019] | -0.55 | -1.31 – 0.21 | 0.154 | -0.44 | -1.46 – 0.58 | 0.401 |
| 2024 [Ref: 2019] | -0.81 | -1.62 – 0.01 | 0.053 | -0.83 | -1.93 – 0.27 | 0.14 |
| (Intercept) | 117.43 ^***^ | 96.14 – 138.71 | **<0.001** | 116.50 ^***^ | 84.21 – 148.79 | **<0.001** |
| **Random Effects** |  |  |  |  |  |  |
| σ^2^ | 3.92 |  |  | 7.05 |  |  |
| τ_00_ | 4.70 _AreaName_ |  |  | 11.01 _AreaName_ |  |  |
| ICC | 0.55 |  |  | 0.61 |  |  |
| N | 148 _AreaName_ |  |  | 148 _AreaName_ |  |  |
| Observations | 1475 |  |  | 1475 |  |  |
| Marginal R^2^ / Conditional R^2^ | 0.665 / 0.848 | |  | 0.640 / 0.842 | |  |
| AIC | 3.92 |  |  | 7.05 |  |  |
| ** p<0.05   ** p<0.01   *** p<0.001* |  |  |  |  |  |  |

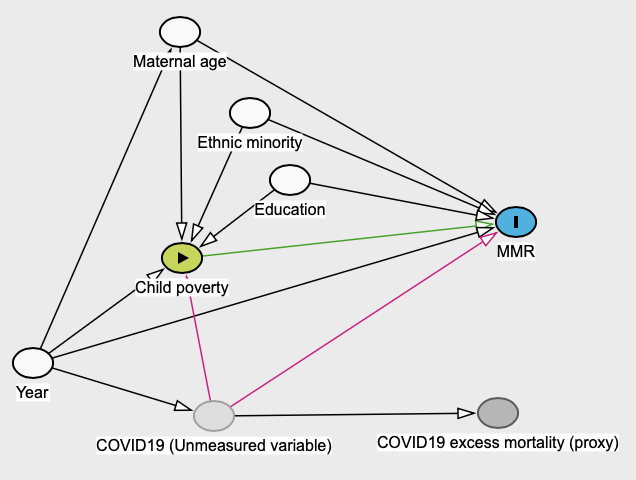

*Figure S1. Logic model identifying key covariates that influence the associations of child poverty on MMR rates.*

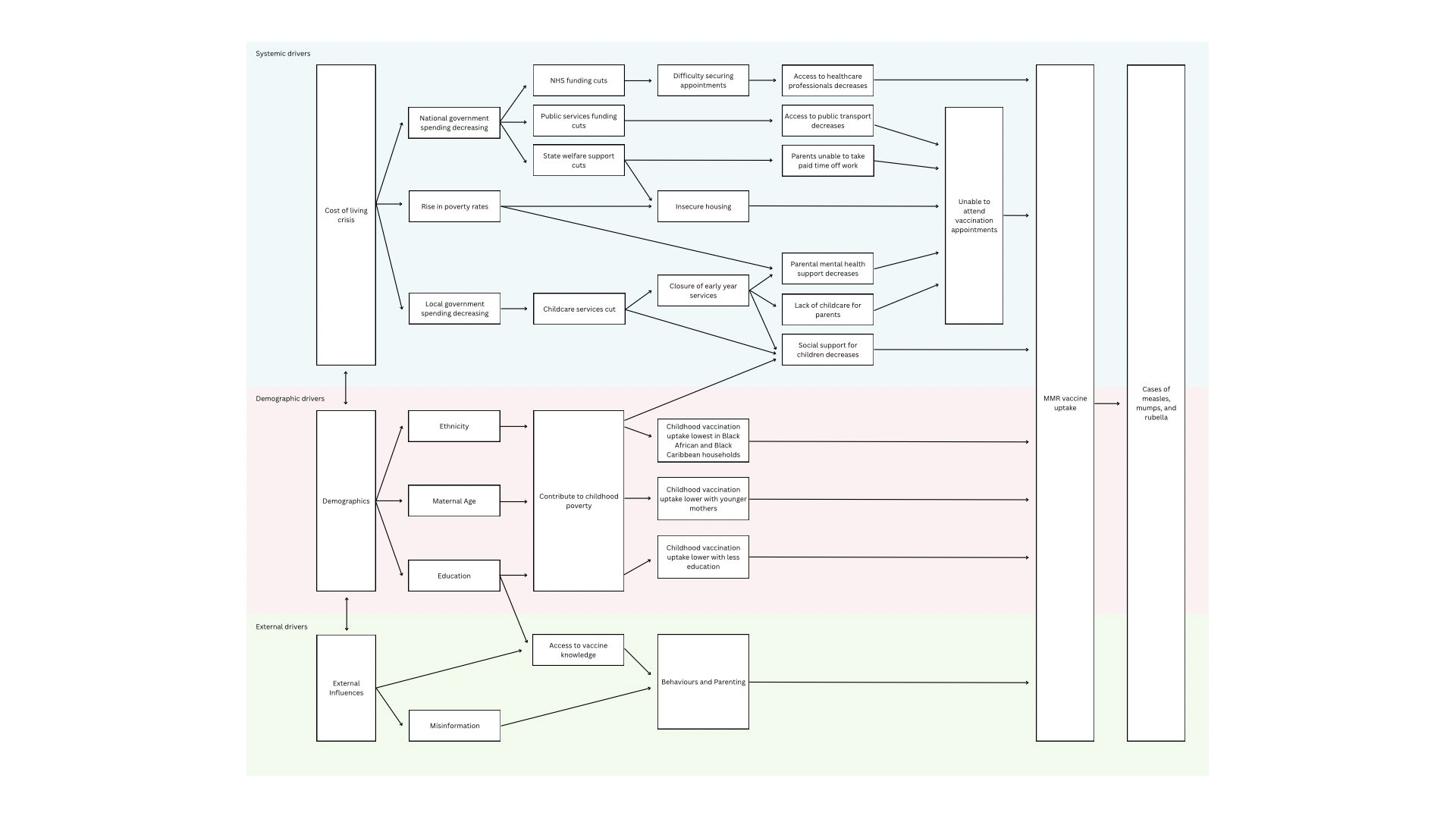

*Figure S2. Logic model illustrating potential pathways from childhood poverty to MMR vaccine uptake with systemic, demographic, and external influences.*

**S.3. Stochastic Modelling of the Impact of Small Reductions in MMR2 Coverage on Measles Transmission**

*S.3.1 Purpose*

This supplementary materials detail the stochastic modelling exercise used to illustrate how a 0.26 percentage‑point reduction in MMR2 vaccination coverage—from 88.8% (the coverage rate in 2015) to 85.5%—affects measles transmission dynamics in populations already below the 95% herd immunity threshold recommended by the World Health Organization (WHO). The model is designed as an illustrative analysis rather than a predictive national forecast, showing why measles transmission is highly sensitive to even small declines in vaccine uptake once population immunity is compromised.

*S.3.2 Rationale for Modelling Approach*

*S.3.2.1 Importance of high MMR2 coverage*

Measles is one of the most infectious pathogens known, with an estimated basic reproduction number (R0) of 12–18, meaning a single infectious case can infect up to 18 susceptible individuals (Robert et al., 2024). Due to this high transmissibility, the WHO recommends ≥95% two‑dose MMR coverage to interrupt sustained transmission (WHO, 2025). Recent UK analyses confirm that England has never achieved this target and that current coverage levels leave large pockets of susceptible children (UKHSA, 2023).

*S.3.2.2 Why a stochastic branching‑process model?*

We chose a Galton–Watson–type branching process because:

1. Measles outbreaks at low coverage are stochastically driven.
2. Branching processes are widely used in UKHSA outbreak nowcasting (Tang et al., 2025).
3. They allow clear visualisation of changes in outbreak size distributions.
4. They align with classical measles modelling for England (Babad et al., 1995).

*S.3.3 Model Structure and Parameters*

A population of 100,000 is assumed, seeded with one infectious case.

Susceptibility is calculated as:

S = 1 − (coverage × vaccine effectiveness)

MMR2 effectiveness is set at 97%, consistent with WHO evidence.

*S.3.3.1*

*Effective reproduction number (Re)*

Re = R0 × S

Using R0 = 15 (Robert et al., 2024), we obtain:

– Baseline (88.8% coverage): Re = 1.965

– Reduced coverage (88.5%): Re = 1.985

This is a ~1.0% increase in effective transmissibility.

*S.3.3.2 Stochastic transmission*

Each infectious individual generates a Poisson(Re) number of secondary infections. Transmission stops when the outbreak dies out or reaches the population cap.

10,000 simulations per scenario were run.

*S.3.4 Results*

*S.3.4.1 Effective reproduction number*

A 0.26 percentage‑point fall in MMR2 coverage yields a 1.0% increase in Re.

Median outbreak sizes:

– Baseline: 34 cases

– Reduced coverage: 36 cases

**This is an 5.9% increase.**

Mean outbreak sizes:

– Baseline: 49 cases

– Reduced coverage: 53 cases

**This is an 8.2% increase.**

Range: 6%–14% increase, depending on whether median or upper‑tail behaviour is emphasised.

*S.3.4.3 Interpretation*

Measles transmission is super‑critical when Re > 1, so small increases in susceptibility amplify outbreak sizes disproportionately.

This is consistent with the 2,911 confirmed measles cases observed in England in 2024 (UKHSA, 2025).

*S.3.5 Limitations*

– Homogeneous mixing assumed.

– No age structure.

– Geographical clustering not modelled.

– Not calibrated to contact matrices.

*S.3.6 Summary and conclusion*

A one percentage point (pp) increase in child poverty rates is associated with a 0.26 pp fall in MMR2 vaccination uptake by the age of 5. Using a stochastic transmission model, a 0.26 percentage‑point fall in MMR2 coverage from 88.8% to 88.5% increases the effective reproduction number (Rₑ) by approximately 1% and increases the expected measles outbreak size by around 6–14%, depending on the metric used. UKHSA reported 2,911 confirmed measles cases in England in 2024 (UKHSA, 2025) such that a mid‑range 10% increase in outbreak size due to increasing poverty rates would correspond to approximately 291 additional confirmed cases (range 175 to 408 additional confirmed cases).

An academic study (Ghebrehewet et al., 2016) estimated that the direct public health costs per confirmed case were £2,714, with additional hospital costs of £1,945 (in 2013 prices). Adjusting these costs using GDP deflators (34)yields an estimated 2024 cost of £6,325 per confirmed case. Applying this updated cost estimate suggests that 291 additional confirmed cases would result in approximately £1.84 million in extra direct and hospital costs (range: £1.12 million to £2.58 million).

Instead of relying solely on confirmed cases, we can consider the wider burden of measles notifications, of which there were 11,162 in 2024. Applying the same mid‑range 10% outbreak‑size increase implies approximately 1,116 additional notifications. The same academic study (5) estimated the cost per notification at £1,790 (2013 prices), equivalent to £2,430 in 2024£. This gives an estimated additional cost of £2.71 million attributable to these excess notifications (range: £1.63 million to £3.80 million).

Between 2015 and 2024, rising child poverty contributed to 42,705 additional 5‑year‑olds who did not receive their MMR2 dose. This accumulation of susceptible children increases the risk and severity of measles transmission. Consistent with this, our stochastic model indicates that a typical poverty‑related 0.26 pp fall in coverage increases the expected outbreak size by around 6–14%, even when baseline coverage is as high as 88.8%.

**References for Supplementary Materials S.3**

Babad, H.R., Nokes, D.J., Gay, N.J., Miller, E., Morgan‑Capner, P. & Anderson, R.M. (1995). Predicting the impact of measles vaccination in England and Wales: model validation and analysis of policy options. Epidemiology and Infection, 114(2), 319–344.

Ghebrehewet S, Thorrington D, Farmer S, Kearney J, Blissett D, McLeod H, et al. The economic cost of measles: Healthcare, public health and societal costs of the 2012–13 outbreak in Merseyside, UK. Vaccine. 2016 Apr;34(15):1823–31. doi:10.1016/j.vaccine.2016.02.029

Robert, A., Suffel, A.M. & Kucharski, A.J. (2024). Long-term waning of vaccine-induced immunity to measles in England: a mathematical modelling study. The Lancet Public Health.

Tang, M.L., McFarlane, I.S., Overton, C.E. et al. (2025). Nowcasting cases and trends during the measles 2023/24 outbreak in England. Journal of Infection.

UK Health Security Agency (2023). Risk assessment for measles resurgence in the UK. [Risk assessment for measles resurgence in the UK](https://assets.publishing.service.gov.uk/media/64aff9448bc29f00132ccd07/risk-assessment-for-measles-resurgence-in-the-UK-2023.pdf)

UK Health Security Agency (2025). Measles: historic confirmed cases, notifications and deaths. [Measles: Historic confirmed cases, notifications and deaths - GOV.UK](https://www.gov.uk/government/publications/measles-historic-confirmed-cases-notifications-and-deaths/measles-historic-confirmed-cases-notifications-and-deaths)

World Health Organization (2025). Critical immunisation thresholds for measles elimination.
